# Paclitaxel Drug Coated Balloon Angioplasty for Medically Refractory Intracranial Atherosclerotic Disease: A U.S. Single-Center Experience with the AGENT balloon

**DOI:** 10.1101/2025.09.17.25336036

**Authors:** Amit Chaudhari, Zachary M. Rosenstein, Rashed Kamal, Mohammed Almajali, Darwin Ramirez-Abreu, Jaafar Kashef Al-Gheeta, Yazan Ashouri, Eugene Lin, Osama O. Zaidat

## Abstract

**Background:** Intracranial atherosclerotic disease (ICAD) is associated with up to 10% of ischemic strokes and a high risk of recurrence. Endovascular treatments including percutaneous transluminal angioplasty and stenting have failed to demonstrate improved outcomes compared to medical therapy alone. Drug coated balloon (DCB) angioplasty has emerged as a promising alternative, though its safety and durability remains uncertain.

**Methods:** This single-center retrospective study analyzed consecutive patients with refractory ICAD treated with AGENT DCB submaximal angioplasty. Patients were treated either for refractory large-vessel occlusion during thrombectomy (emergency rescue) or for recurrent ischemic symptoms from high-grade stenosis (elective primary). Technical success was defined as <50% residual stenosis without adjunctive stenting. Safety outcomes included hemorrhage, dissection, stroke, and death at three months.

**Results:** Of the 11 identified patients, nine underwent successful DCB angioplasty, five for emergent rescue therapy and four for elective primary therapy. There was a combined 78% technical success rate, with no major procedural complications. Mean stenosis reduction was 53.6% (paired Wilcoxon p=0.014; pre-procedure mean (SD) stenosis 90.8% (±8.6%) to post-procedure 37.3% (±33.4%)). Restenosis occurred in three of the four (75%) elective primary patients without recurrent ischemic events. Follow-up angiographic data for the emergent rescue cohort was unavailable, though no symptomatic ischemic events were reported.

**Conclusion:** Paclitaxel-coated AGENT drug-coated balloon (DCB) submaximal angioplasty was safe and yielded immediate improvements in luminal diameter. However, high rates of re-stenosis emphasize the need for innovative devices and larger prospective studies to define its therapeutic role for refractory ICAD.

## Introduction

Intracranial atherosclerotic disease (ICAD) is associated with up to 10% of ischemic strokes and is associated with a high risk of recurrence. The landmark SAMMPRIS and VISSIT randomized clinical trials demonstrated that percutaneous transluminal angioplasty and stenting (PTAS) did not improve outcomes compared to maximum medical therapy alone.^1–3^ Early data with paclitaxel DCBs, such as SeQuent please and Taijieweiye, show technical feasibility, though optimal dosing, procedural technique, and long-term outcomes remain unclear.^4^ Moreover, neither device is commercially available in the U.S, thus limiting the evidence-based application of this strategy in clinical practice for the U.S. population.

Here, we present the first U.S. single-center therapeutic experience with the paclitaxel-coated AGENT DCB (Boston Scientific, Marlborough, MA, U.S.) for submaximal angioplasty of symptomatic ICAD refractory to maximal medical therapy, highlighting technical performance, safety profile, and clinical outcomes. The analysis is stratified by indication, emergent rescue angioplasty during failed mechanical thrombectomy vs. elective primary angioplasty for recurrently symptomatic patients despite maximal medical therapy, to inform the design of future prospective investigations.

## Methods

### Study Population

Institutional review board approval was obtained for this study, with a waiver of informed consent for the retrospective review of de-identified clinical data. We reviewed all consecutive intracranial DCB angioplasties performed at our tertiary care center between January 2025, and July 2025. Eligible patients were adults (≥18 years) with symptomatic ICAD who underwent submaximal angioplasty for either: (1) refractory large-vessel occlusion during mechanical thrombectomy, defined as failure to achieve recanalization after ≥3 device passes or immediate re-occlusion (emergent rescue cohort); or (2) high-grade (>70%) stenosis with recurrent ischemic symptoms despite maximal medical therapy (elective primary cohort). Patients for whom DCB angioplasty was selected but the balloon could not be advanced technically to the target lesion were excluded from the final analysis.

### Procedure

All procedures were performed under general anesthesia by experienced neurointerventionalists using biplane digital subtraction angiography. All patients were pre-treated with dual antiplatelet therapy and systemic anticoagulation with intravenous heparin was administered to maintain an activated clotting time (ACT) of 250–300 seconds throughout the procedure.

Conventional techniques used for drug-coated balloon (DCB) angioplasty, particularly with devices such as the SeQuent Please, often involve vessel pre-dilation with an uncoated balloon to achieve a residual stenosis of less than 50% prior to the application of the DCB.^5–7^ In the present study, we deemed this unnecessary as robust evidence from multicenter randomized trials for coronary DCB angioplasty suggests that the AGENT™ DCB is able to successfully dilate the affected stenotic vessel primarily.^8^ As such, an AGENT DCB selected to match approximately 80% of anticipated parent vessel diameter was advanced primarily over the microwire to span the entire stenotic lesion. Inflation was performed to nominal pressure for 60 seconds to allow uniform drug transfer to the vessel wall (see Figure 1). If significant residual stenosis (>50%) persisted, a second prolonged inflation was attempted at the neurointerventionalist’s discretion.

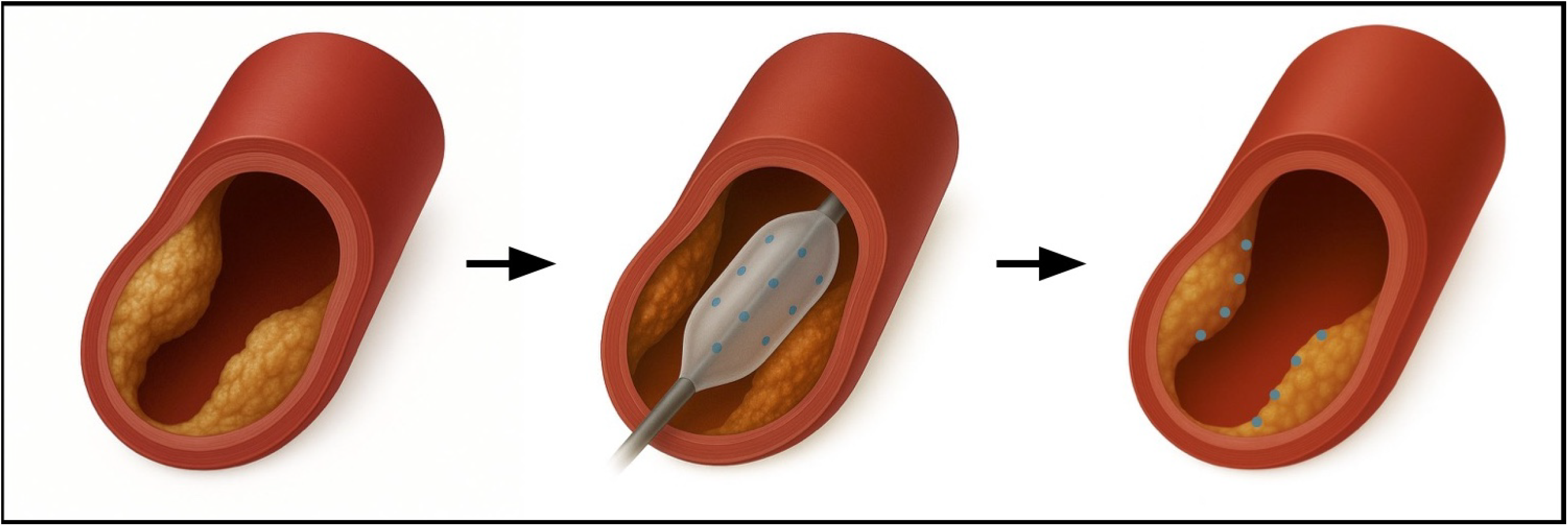

Following submaximal angioplasty, repeat angiography was performed to evaluate the residual stenosis, flow, and potential complications, including dissection or vessel rupture. Intracranial stenting was reserved for cases of inadequate angiographic result, flow-limiting dissection, or elastic recoil after balloon angioplasty. Post-procedural antiplatelet therapy consisted of dual antiplatelet therapy with aspirin and a P2Y12 inhibitor, continued for a duration specified per institutional protocol.

### Data Collection and Outcomes

Clinical and procedural data were abstracted from electronic medical records and angiographic archives. Lesion characteristics, procedural details (balloon size, inflation time), and use of adjunctive devices were recorded. Stenosis severity was quantified via digital subtraction angiography by the treating neurointerventionalist according to the Warfarin-Aspirin Symptomatic Intracranial Disease (WASID) criteria, both before and after AGENT DCB submaximal angioplasty. The primary outcome was technical success, defined as <50% residual stenosis after DCB angioplasty without the need for adjunctive PTAS. Safety outcomes included periprocedural intracranial hemorrhage, vessel dissection, symptomatic re-occlusion, ischemic stroke, and mortality at 1 month and at 3 months.

### Statistical Analysis

Given the small cohort size, analyses were primarily descriptive. Continuous variables are presented as mean ± standard deviation, while categorical variables are reported as counts and percentages. The within-patient change in stenosis severity was assessed using the paired Wilcoxon signed-rank test. All statistical tests were two-sided with an alpha of 0.05. P-values are considered exploratory and were not adjusted for multiplicity.

## Results

### Patient Characteristics

A total of 11 consecutive patients (5 female, 45%) with medically-refractory ICAD were identified as suitable candidates for AGENT DCB submaximal angioplasty. Two patients initially selected for DCB angioplasty were excluded intraprocedurally because of technical inability to advance the AGENT DCB to the target lesion due to proximal vessel tortuosity. Eight out of the remaining nine patients had hypertension (88.9%), with other prevalent risk factors including hyperlipidemia (88.9%) and diabetes mellitus (33.3%). The pre-procedure National Institutes of Health Stroke Scale (NIHSS) score was variable with mean (±SD) of 6.6 (±8.2). Further demographical data is presented in Table 1.

**Table 1:** Pre-Procedural and Procedural Characteristics. **Table 1. Baseline Demographics and Clinical Characteristics**. Data are presented for the entire patient cohort (All) and stratified by procedural group (emergency rescue vs. elective primary).

| Variables | All (n = 9) | Rescue (n = 5) | Primary (n = 4) |
| --- | --- | --- | --- |
| **Demographics** |  |  |  |
| Male, n (%) | 4 (44.4%) | 3 | 1 |
| Age, mean ± SD (y) | 63.1 ± 13.3 | 69.0 ± 13.9 | 55.8 ± 9.3 |
| Race |  |  |  |
| White | 8 (88.9%) | 5 | 3 |
| Hispanic | 1 (11.1%) | 0 | 1 |
| **Past Medical History** |  |  |  |
| Diabetes Mellitus | 3 (33.3%) | 1 | 2 |
| Hypertension | 8 (88.9%) | 5 | 3 |
| Atrial Fibrillation | 3 (33.3%) | 2 | 1 |
| Hyperlipidemia | 8 (88.9%) | 4 | 4 |
| Congestive Heart Failure | 1 (11.1%) | 1 | 0 |
| Prior stroke | 6 (66.7%) | 3 | 3 |
| Prior MI | 2 (22.2%) | 2 | 0 |
| Smoking | 4 (44.4%) | 2 | 2 |
| BMI, (kg m<sup>-2</sup>) | 33.3 ± 7.6 | 34.6 ± 10.0 | 31.6 ± 3.7 |
| CKD | 0 (0%) | 0 | 0 |
| **Presentation** |  |  |  |
| NIHSS Score, mean (SD) | 6.6 ± 8.2 | 11.4 ± 8.2 | 0.5 ± 1.0 |
| **Lesion Description** |  |  |  |
| Mean pre-procedural stenosis | 90.8 ± 8.9% | 92.0 ± 8.4% | 89.3 ± 10.5% |
| ICA | 2 (22.2%) | 0 | 2 |
| MCA | 4 (44.4%) | 3 | 1 |
| PCA | 1 (11.1%) | 1 | 0 |
| Vertebrobasilar | 2 (22.2%) | 1 | 1 |

*Abbreviations: BMI, body mass index; CKD, chronic kidney disease; MI, myocardial infarction; n, number of participants; NIHSS, National Institutes of Health Stroke Scale; NS, not significant; SD, standard deviation; TTE, transthoracic echocardiogram.*

Seven of the eleven selected patients presented with acute ischemic stroke and underwent mechanical thrombectomy with failure to achieve recanalization after ≥3 device passes or immediate re-occlusion (emergent rescue cohort). Two patients from this emergency rescue cohort were excluded from the analysis because of technical inability to advance the AGENT DCB to the target lesion due to proximal vessel tortuosity. The remaining five patients underwent DCB angioplasty, three to the middle cerebral artery (MCA), one to the posterior cerebral artery (PCA), and one to the mid-basilar artery (BA).

Four of the eleven patients were selected for high-grade (>70%) stenotic lesions with recurrent ischemic symptoms despite maximal medical therapy (elective primary cohort). All four patients underwent DCB angioplasty, two to the distal intracranial internal cerebral artery (ICA), one to the MCA, and one to the mid-BA.

The mean pre-procedural stenosis, as measured by the WASID criteria, was not significantly different between the two cohorts (mean (SD) of stenosis 92.0 (±8.4%) in emergent rescue cohort and 89.2% (±10.5%) in the elective primary cohort).

### Safety Outcomes

AGENT DCB submaximal angioplasty was very well tolerated in this patient population. Three patients in the emergency rescue cohort had small asymptomatic petechial hemorrhages noted on the follow-up MRI, all classified as Hemorrhagic Infarction Type 1 (HI-1) on the Heidelberg Bleeding Classification, thought to be related to natural stroke progression rather than the DCB angioplasty procedure.^9^ No major symptomatic bleeding events were observed. There were no reports of peri-procedural vessel dissections, symptomatic re-occlusions, ischemic strokes in either cohort within 30 days of the intervention.

### Efficacy Outcomes

Nine of the eleven selected patients underwent successful AGENT DCB submaximal angioplasty, with significant improvements in luminal diameter, with a mean absolute reduction of 53.6% (paired Wilcoxon p = 0.014; pre-procedure mean (SD) stenosis 90.8% (±8.6%) to post-procedure 37.2% (±33.4%)). Efficacy of the intervention was not significantly different between the two cohorts (see Figure 2). Seven of the total nine patients (78%) met the primary outcome for technical success with <50% residual stenosis, with only two patients, both in the emergency rescue cohort, requiring rescue PTAS for persistent flow-limiting stenosis despite DCB angioplasty to the middle cerebral artery.

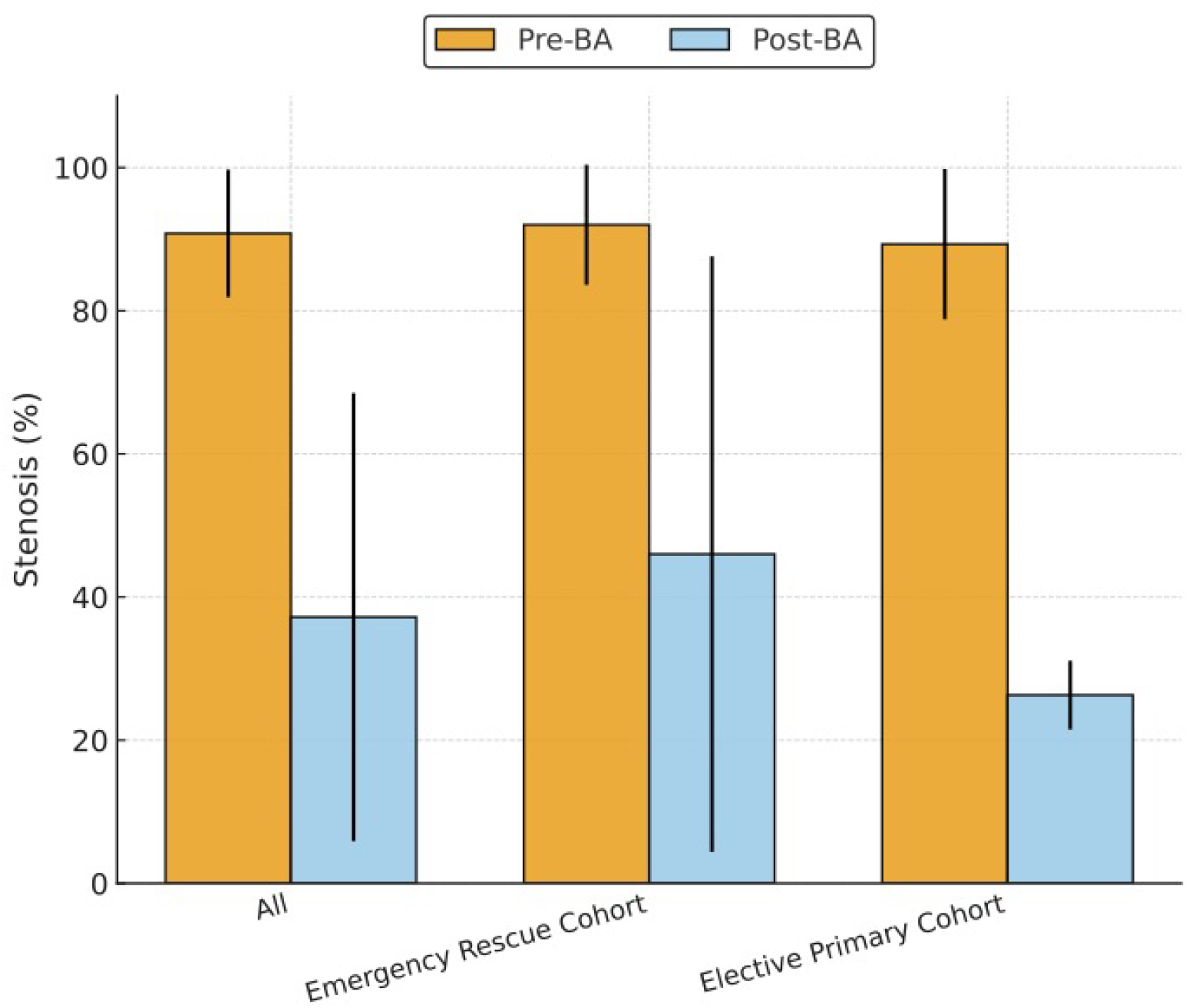

### Follow-up Outcomes

There was limited 3-month angiographic follow-up data to assess for delayed asymptomatic re-stenosis, with none of the patients in the emergent rescue therapy cohort obtaining repeat imaging as of yet. All four patients in the elective primary cohort did receive follow-up non-invasive angiographic imaging (mean follow-up 64 days), with three of the four (75%) patients showing recurrence of the intracranial stenosis but without clinical symptomatic ischemic events.

There were no incidences of procedure-related mortality in either of the cohorts within the 3 month follow-up period. However, three patients, again all in the emergency rescue cohort, did die from non-procedure-related causes, including one from gastrointestinal bleed and pneumonia leading to refractory septic shock, one from goals of care discussions due to completed bilateral cerebellar and brainstem ischemic stroke, and one from self-inflicted asphyxiation as part of a suicide attempt.

## Discussion

This single-center U.S. experience demonstrates that submaximal angioplasty with the commercially-available paclitaxel-coated AGENT drug-coated balloon (DCB) is technically feasible, safe, and effective in reducing intraluminal stenosis in patients with symptomatic intracranial atherosclerotic disease (ICAD) refractory to maximum medical therapy. Our results show a combined 78% technical success rate, with no major periprocedural complications and low need for adjunctive stenting. These findings add to the limited body of literature on DCB angioplasty for ICAD, particularly in the United States where the SeQuent Please DCB and Taijieweiye DCB used in the existing literature are not commercially available.

Intracranial atherosclerotic disease (ICAD) accounts for up to 10% of ischemic strokes and is associated with a high risk of recurrence.^10^ The landmark SAMMPRIS and VISSIT randomized clinical trials demonstrated that percutaneous transluminal angioplasty and stenting (PTAS) did not improve outcomes compared to maximum medical therapy alone.^1–3^ These findings limited the use of PTAS, but left a therapeutic gap for patients with ICAD refractory to medical therapy. Alternative strategies including primary balloon angioplasty alone have been attempted, but with limited success and high periprocedural complications.^11^

Drug-coated balloon (DCB) angioplasty has emerged as a promising approach, delivering localized antiproliferative therapy while minimizing the risks of in-stent restenosis and delayed thrombosis associated with permanent scaffolds.^12,13^ Early reports using paclitaxel-coated DEBs, particularly the SeQuent Please resveratrol-coated paclitaxel balloon (SeQuent Please; B. Braun, Berlin, Germany), have demonstrated technical feasibility and short-term angiographic improvement.^14,15^ This is further being investigated in the ongoing multicenter randomized Dr. BEYOND trial, which compares the efficacy and safety of DCB angioplasty with the paclitaxel-coated Taijieweiye DCB to bare-metal stenting with the the Wingspan stent system.^16^ Nonetheless, current literature remains sparse, with optimal drug dose, coating excipient, procedural workflow, and long-term outcomes remaining poorly defined. Moreover, neither the SeQuent Please DEB nor the Taijieweiye DCB are available for commercial use in the United States (U.S.), thus limiting safety and efficacy investigations and evidence-based application of this strategy in U.S. clinical practice.

AGENT (Boston Scientific, Marlborough, MA, U.S.) is an FDA-approved coronary DCB commercially available in the U.S., with multicenter randomized trials demonstrating non-inferiority compared to the SeQuent Please for in-stent coronary stenosis at six months and superiority against uncoated balloons in the AGENT IDE trial.^8,17^ Unlike SeQuent Please, it employs a low-dose (2 µg mm^-2^) crystalline paclitaxel coating with an acetyl-tributyl-citrate excipient, and a laser-bonded tip and bi-segment shaft designed to enhance navigability in tortuous anatomy. While widely used in the coronary circulation, the AGENT DCB has unique delivery and pharmacokinetic characteristics which have not been systematically studied for intracranial use.

Our series provides early insight into the use of the paclitazel-coated AGENT DCB for submaximal angioplasty of symptomatic ICAD refractory to maximal medical therapy. We describe 11 patient cases, seven as emergent rescue therapy for refractory large-vessel occlusion during mechanical thrombectomy and four as elective primary therapy for high-grade (>70%) stenosis with recurrent ischemic symptoms despite maximal medical therapy. Only two of the 11 DCB angioplasty attempts were unsuccessful, both due to the highly tortuous anatomy of their proximal vessels. Newer device interactions specifically designed for intracranial use may improve the maneuverability and deliverability of DCBs to avoid such events in the future.

Nine patients underwent successful intracranial AGENT DCB submaximal angioplasty, five as emergency rescue therapy and four as elective primary therapy. Safety outcomes were favorable with no peri-procedural major hemorrhages, vessel dissections, symptomatic re-occlusions, ischemic strokes in either cohort within the first month. Petechial hemorrhages observed in three patients within the emergent rescue cohort were asymptomatic, classified HH-1 on the Heidelberg Bleeding Classification, and consistent with hemorrhagic transformation of ischemic stroke rather than a procedural complication. Nonetheless, multiple manipulations of the vascular endothelium can induce injury and, combined with dual antiplatelets and systemic heparinization, can increase the risk of vessel dissection or rupture.^18,19^ Mortality events were unrelated to the procedure.

Technical efficacy of the procedure was acceptable in the short-term. There were significant reductions in intraluminal narrowing to <50% residual stenosis in seven of the nine patients (78%). Two patients, both undergoing emergency rescue therapy, remained refractory and required rescue PTAS, comparable to the failure rates described in current literature.^20^ Both had MCA stenosis, and failure of DCB angioplasty was likely mediated by the patient’s individual pathology rather than the location of the stenosed vessel. Of the successful angioplasties, there were no reports of symptomatic re-occlusions or ischemic strokes within three months.

However, the durability of DCB angioplasty remains a concern. Three of the four elective patients in our cohort demonstrated restenosis at short-term follow-up despite dual antiplatelet therapy. This finding echoes prior experience with angioplasty and underscores the need to temper initial enthusiasm: while DCB angioplasty may serve as a safe temporizing strategy, it is unlikely to represent a definitive solution for refractory ICAD where stenotic lesions are prone to recoil and re-stenosis.^21^ Chen and colleagues have identified predictors of restenosis after DCB treatment.^22^ Meanwhile, Xue and colleagues reported 100% technical success with no restenosis when treating intracranial in-stent restenosis.^23^ In contrast, the universal restenosis in our primary cohort suggests that *de novo* atherosclerotic plaques may have a more aggressive biology.^24^ DCBs designed specifically for the unique mechanical and biological environment of intracranial arteries may be able to achieve durable results. Coating with medications other than paclitaxel, such as sirolimus, may also further improve long-term efficacy.^25^ Future research should investigate the optimal medication, dosing, duration of endothelial contact, and other technical technical details that could make the procedure more durable.

Our findings provide a proof-of-concept for the use of AGENT DCB submaximal angioplasty for symptomatic refractory ICAD to inform clinical practice and future device development. The study is understandably limited by its small sample size, retrospective single-center design, heterogeneous patient population, and lack of core-lab adjudication of angiographic measurements. Procedurally, unlike most international studies with SeQuent Please, we did not perform predilation with a conventional balloon.^26^ Based on cardiac literature demonstrating the efficacy of the semi-complaint AGENT DCB in reducing coronary stenosis, we believe that the semicompliant AGENT platform provides sufficient submaximal angioplasty without predilation along with benefits of delivering the paclitaxel medication to an uninterrupted endothelium throughout the vascular remodeling process, though this assumption warrants further evaluation in ICAD. Lastly, the study had limited angiographic follow-up and even this was incomplete, particularly in the emergent thrombectomy group, preventing systematic assessment of asymptomatic re-stenosis long-term.

To our knowledge, this is the first evidence-based case series systematically investigating the use of paclitaxel-coated AGENT DCB submaximal angioplasty as an off-label alternative for patients with symptomatic refractory ICAD, particularly in the U.S. where commercial availability of DCBs like SeQuent Please and Taijieweiye remains uncertain. The study shows significant immediate improvements in intraluminal stenosis with low peri-procedural complication rates. However, the high rate of restenosis in the elective primary cohort underscores that this approach is best considered a temporizing measure rather than a definitive solution. Future prospective, multicenter studies, such as DR. BEYOND, are essential to define the long-term efficacy of DCB angioplasty and to guide the design of next-generation devices tailored specifically for the intracranial circulation with an emphasis on patient-specific factors like lesion morphology and inflammatory state. Until such evidence is available, DCB angioplasty should be considered a salvage strategy rather than a first-line intervention.

## Conclusion

Paclitaxel-coated AGENT drug-coated balloon (DCB) submaximal angioplasty was safe and produced significant immediate improvements in luminal diameter for patients with symptomatic ICAD refractory to maximum medical therapy. However, the effect was short-lived and a significant number of patients experienced re-stenosis, though without any reports of recurrent ischemic events. Taken together, these findings suggest that DCB angioplasty is a feasible temporizing option in carefully selected patients but should not replace best medical management as first-line therapy. Larger, prospective studies are required to clarify the safety, durability, and ultimate role of this emerging strategy.

## Data Availability

The data supporting the findings of this study are available within the article. Further details are not publicly available due to patient privacy restrictions, but can be provided by the corresponding author upon reasonable request.

